# Sequential informed pooling approach to detect SARS-CoV2 infection

**DOI:** 10.1101/2020.04.24.20077966

**Authors:** Renato Millioni, Cinzia Mortarino

## Abstract

The alarming spread of the pandemic coronavirus disease 2019 (COVID-19) caused by the SARS-CoV-2 virus is requiring several measures to reduce the risk of contagion. Every successful strategy in controlling SARS-CoV2 infection depends on the timely viral diagnosis which should include asymptomatic carriers. Consequently, strategies to increase the throughput for clinical laboratories to conduct large-scale diagnostic testing are urgently needed. Here we support the hypothesis that standard diagnostic protocol for SARS-CoV-2 virus could be conveniently applied to pooled samples obtained from different subjects. We suggest that a two-step sequential pooling procedure could identify positive subjects, ensuring at the same time significant benefits of costs and time. Simulation data are used to assess the efficiency, in terms of number of required tests, both for random assignment of the subjects to the pools and for situations when epidemiological and clinical data are used to create an “informed” version of the pooling. Different scenarios are examined in the simulations to measure the effect of different pool sizes and different values for the virus frequency. Our results allow to customize the pooling strategy according to the specific characteristics of the cohort to be tested.

## Introduction

The pandemic coronavirus disease 2019 (COVID-19) is caused by the SARS-CoV-2 virus. The infection is predominantly transmitted through large droplets and by contact with infected surfaces or fomites. The alarming spread of the infection and the severe clinical disease that it may cause have led to several measures to reduce the risk of contagion. Active case detection, rapid case isolation and contact quarantine, as well as rigorous application of infection control practices are successful strategies in controlling SARS-CoV2 infection outbreaks. The first step on which these strategies are set is the viral diagnosis. The overloading to which the laboratories are currently subjected causes a cascade delay of all virus containment procedures with potential dramatic results for the prevention of the infection.

In most countries, testing for COVID-19 is mainly restricted to people with symptoms. However, a large percentage of asymptomatic subjects is estimated to exist. Asymptomatic spread has likely driven the silent growth of SARS-CoV2 epidemic which emerged only when the health system began to collapse. Asymptomatic cases play a role in the transmission and thus pose a significant infection control challenge. How much asymptomatic individuals affect the virus diffusion is actually considered a crucial task to evaluate [1]. Tracing contacts of known positive cases, travel bans and social distancing are the main strategies to reduce the risk of contagion due to asymptomatic subjects. A widespread testing strategy to screen asymptomatic subjects could be useful in reducing transmission of SARS-CoV2, but this approach is highly challenging taking into account of the amount of work, time and costs that it would entail.

For this reason, we propose here a pre-screening strategy which should increase the capacity for clinical laboratories to conduct large-scale diagnostic testing, enough to screen a significant portion of the asymptomatic population.

SARS-CoV2 is an enveloped viruses containing a single strand of positive-sense RNA and its diagnostic protocol is a RT-PCR assay, as previously described in details [2, 3]. Briefly, SARS-CoV2 have been detected from a variety of upper and lower respiratory sources including throat, nasal nasopharyngeal (NP), sputum, and bronchial fluid [4, 5]. Oropharyngeal (OP) and NP swabs are the most frequently used samples. The sampling is carried out using two distinct swabs which can be inserted in the same test tube containing the viral transport medium to increase the yield for the RT-PCR analysis [6]. Total RNA is extracted and SARS-CoV2 target genes are simultaneously amplified and tested during the quantitative RT-PCR assay.

Recently, Hogan et al (2020) [7] performed a retrospective study on SARS-CoV-2 based on sample pooling. “Pooling” means that swab samples taken from different subjects can be combined before the RNA extraction phase. These Authors used 2888 samples from nasopharyngeal and bronchoalveolar lavages that were collected between January 1, 2020, and February 26, 2020, from subjects who had not been tested for SARS-CoV-2. Nine or ten samples were pooled, and screening was performed by RT-PCR. A total of 292 pools were screened and the confirmed positivity rate for SARS-CoV-2 was 0.07% (2/2888). The aim of pooling is to reduce the number of test kits used, significantly shortening the time and costs of analysis. As reported by on-line media (https://www.tabletmag.com/sections/science/articles/pooling-covid-19-israel), a pooling approach has also recently being successfully used in Israel but, to the best of our knowledge, no studies have been published yet. Scientists from the Technion Israel Institute of Technology stated that when 64 samples are mixed together, the analysis on the pool test gives a positive result even if just one of the swabs came from an infected subject. The pooled sample can give laboratory results of equal quality of those with individual testing because there are no changes to the RT-PCR diagnostic protocol.

The basic concepts for understanding the pooling strategy are simple: 1) only a pool made up of all negative samples will give a negative result for the pool analysis; 2) a single positive sample within a pool makes the result of the pool analysis positive. If the pool is positive, it is necessary to proceed to individual testing, to identify the true positives (TP) and the false positives (FP: i.e., a negative subject whose swab has been mixed with at least one positive swab). As all individual samples in a negative pool are considered as true negative (TN), the pooling approach significantly reduces time and cost when a large proportion of pools tests negative. However, it is clear that the effectiveness of pooling is inversely proportional to the frequency of the virus in the selected cohort and, as we will demonstrate more precisely in the results section, this approach can be inefficient or even counter-productive if the presence of the virus is high.

The aim of this paper is to i) propose a two-step sequential pooling strategy, ii) identify the variables for which the pooling method can be more or less effective and to iii) develop strategies to further improve this approach. For this purpose, we began by identifying the main variables which should be included in our model. The first and perhaps most important variable, as already mentioned, is the frequency of the virus. Unfortunately, this information is not known *a priori* but it can be estimated. The second variable is the effectiveness of the clinical and epidemiological criteria that can be adopted to create the pools, compared to an analysis in which these pools are created randomly. The third variable is the size of the pool. We have taken into consideration a wide range of scenarios to adjust the variables to give the best result with fewer tests.

As pooling strategies that can improve the pooling approach, we compared alternative methods to create pools and evaluate their different performances according to the different conditions of the variables previously described. Our data suggest that pre-screening strategy based on the use of a sequential informed pooling approach ensures that, in the most favourable conditions with low virus frequency, the number of tests can drop to 20% of the number of test required by individual testing. Higher virus frequencies still make sequential pooling efficient, provided pool size is decreased and/or reliable epidemiological and clinical data are used to create pools.

## Methods

In the proposed procedure, for each patient involved in the study, three distinct swabs will be performed, following the standard protocols. The first and second swab will be used for the creation of the pools that we will define below. The third swab will be kept and eventually used as a validation test.

The swab used for the creation of a pool “H” (sample 1) is firstly placed in a single sample tube and subsequently transferred in a pooled sample tube together with the sample 1 of other subjects (Figure 1). It is important that the pooled sample could be analyzed with the same standard procedures that are applied for individual samples.

**Figure 1.**
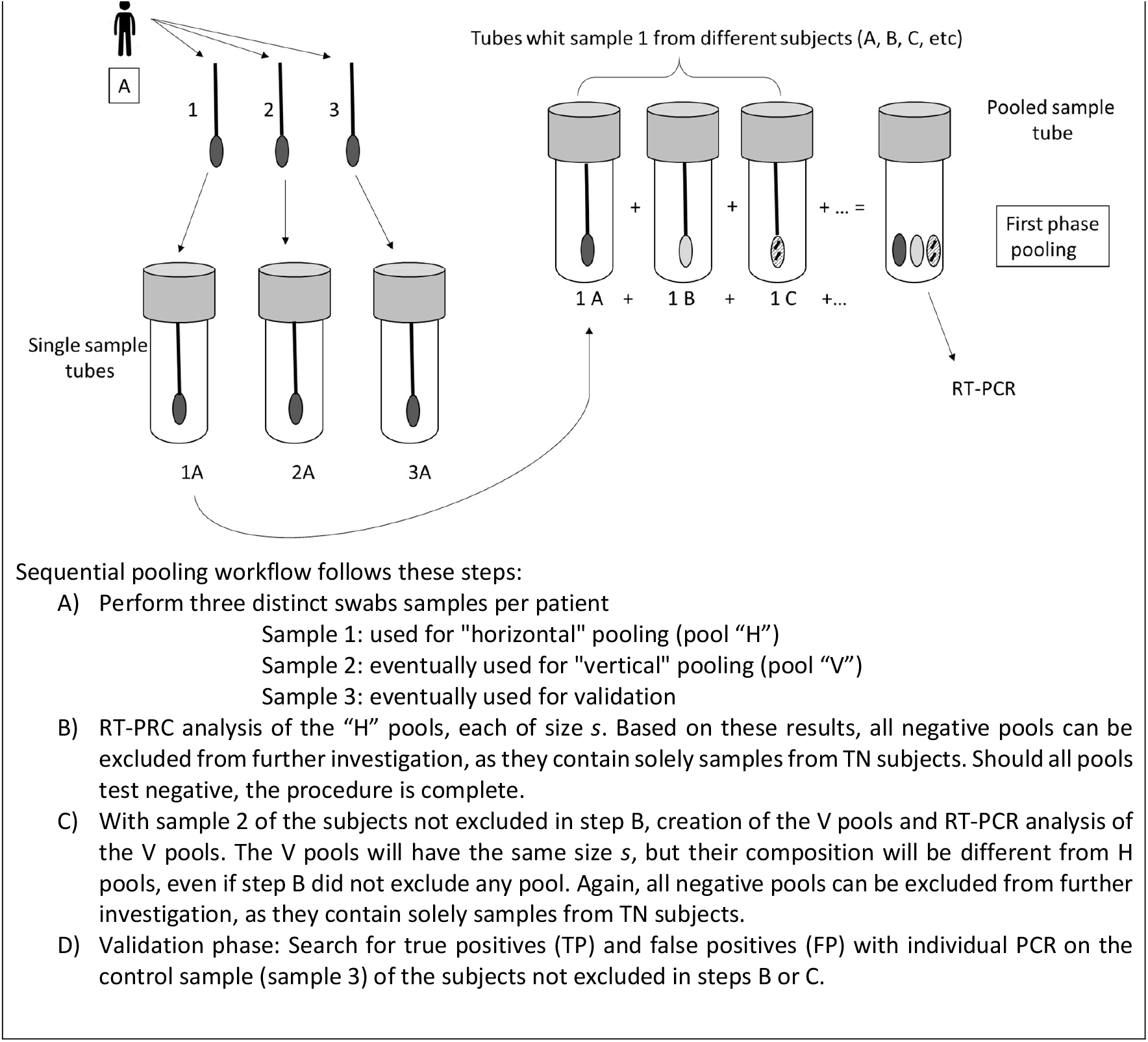
Sequential pooling workflow.

The informed sequential pooling follows exactly the same procedure with the only difference that a score about the probability to be infected will be associated to each subject, in order to tag the subject as “suspected positive” or “suspected negative”. The aim is to include in the same pool subjects with the higher scores, avoiding their random spreading in the matrix. The correct assignment of this score should be accomplished by compiling a dedicated questionnaire. The score is calculated on the basis of clinical and epidemiological criteria that have already been associated to a higher risk of acquiring COVID-19 [8]. For instance, susceptibility seems to be strongly associated with age and biological sex [9, 10, 11] suggesting that these simple criteria may play a pivotal role for the pool assignment.

In Figures 2 and 3, we show a simple graphic representation of the sequential pooling and of the informed sequential pooling approach respectively. For these explicative images, we have chosen a cohort to test of dimension *N* equal to 30, just to facilitate the visual representation.

**Figure 2.**
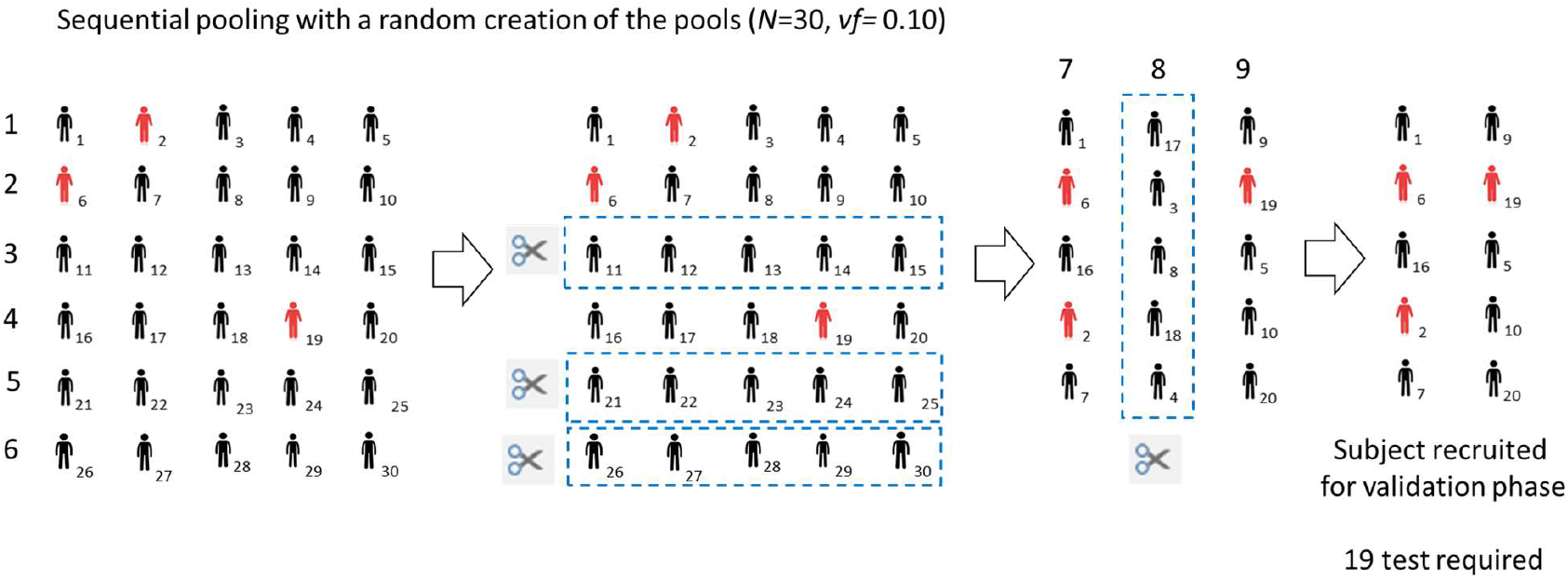
Graphic representation of the sequential pooling approach. The cohort dimension is *N*=30, the pool size is 5, the virus frequency, *vf*, is 0.10.

**Figure 3.**
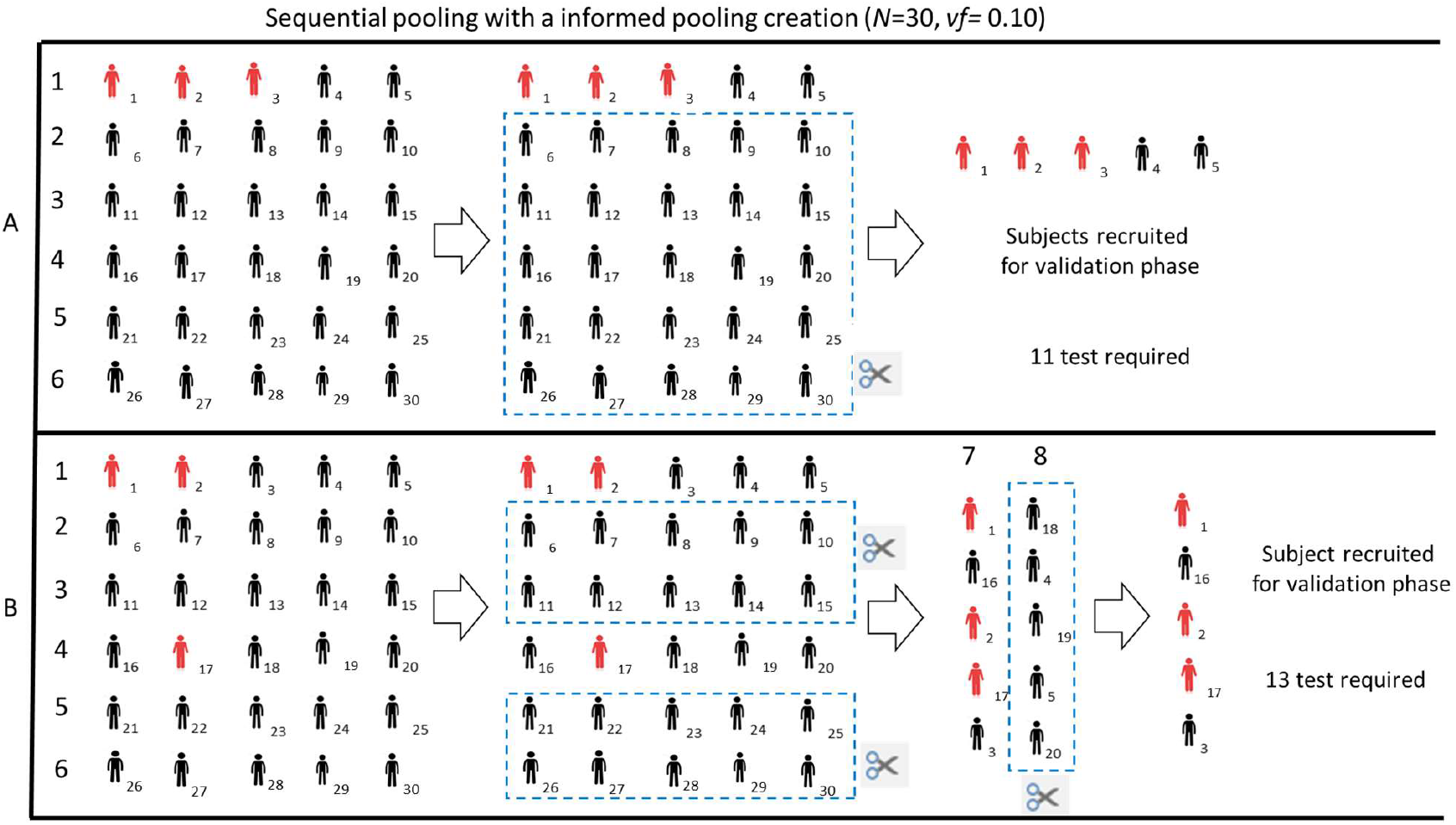
Graphic representation of the sequential informed pooling approach. The cohort dimension is *N*=30, the pool size is 5, the *vf* is 0.10. Panel A and B show two possible scenarios according to a different reliability of the information available to classify subjects as “suspect positive” or “suspect negative”.

In Figure 3, the upper panel shows a hypothetical scenario for which all positive subjects are grouped in the first pool. This result can be obtained if the information available to classify subjects as “suspect positive” or “suspect negative” are optimal. In the lower panel, we show another scenario for which clinical and epidemiological information allowed a grouping of the positives subjects which is only partially correct.

However, also in this case, the informed approach is still useful to improve the efficiency of the method as compared to a random creation of pools.

## Results

In order to assess the advantage of this two-step sequential pooling strategy in comparison with a standard approach, where each subject’ swab is separately tested, we performed simulations^1^ under different conditions. We assumed to analyse a group of *N*=600 subjects. The size *s* of each pool (both H and V) was allowed to vary from 2 to 300.^2^ We examined a virus frequency, *vf*, ranging from 0.01 to 0.30 (the latter situation thus corresponding to 30% of the subjects TP to the virus).

Preliminary, we examined the performance of this strategy without using prior information about the subjects, that is creating pools completely at random. To do this, after setting *s* and *vf*, we performed 5,000 simulations and we recorded the ratio between the total number of swab tests required, *T*, and *N*. For two-step sequential pooling, *T* includes both H and V pools required in steps B and C, but also validation tests in step D on all the swabs from subjects not previously excluded. Since, without a pooling strategy, *N* tests should be performed, the ratio *T/N* measures the efficacy of the proposed procedure. The smaller is its value, the larger is the reduction of required tests. Conversely, values close to 1 (or even above 1) would represent a useless (or a counter-productive) strategy. Table 1 and Figure 4 show the results for *s* equal to 5, 12 and 24 (the entire set of plots is available in the Supplementary Material section, Figure S.1). The curves plotted represent the 1st, the 25th, the 50th (median), the 75th, and the 99th percentiles of *T/N* obtained in the set of 5,000 simulations, for different *vf* values. In particular, the 1st and the 99th percentiles give an idea of the range of *T/N* between “lucky” or “unlucky” assignments to the pools. The spread between the 25th and the 75th, which is always very small in Figure 4, represents the central half of the simulations (after excluding the 25% more “lucky” and the 25% more “unlucky” ones). As the pool size increases, we notice that the curves are less linear and the spread between the 1st and the 99th percentile increases. For very small pools (*s*=3), with a low virus frequency, the number of tests required is about 40% with our procedure as compared to separately testing each subject. As *vf* moves to larger values, the number of tests grows slowly and the pooling is still efficient (*T/N*<1) even if 25% of the subjects are positive in the group. Conversely, if we use larger pools (*s*=24), the number of tests could drop to 20% for low virus frequency. However, the number of tests would increase faster as *vf* grows, and the procedure would be efficient only up to about 10% of positive subjects in the analysed cohort. In summary, the linear path of small pools ensures efficiency even for larger *vf*, but the nonlinear path of larger pools allows great efficiency for populations with a low virus presence.

**Table 1.**
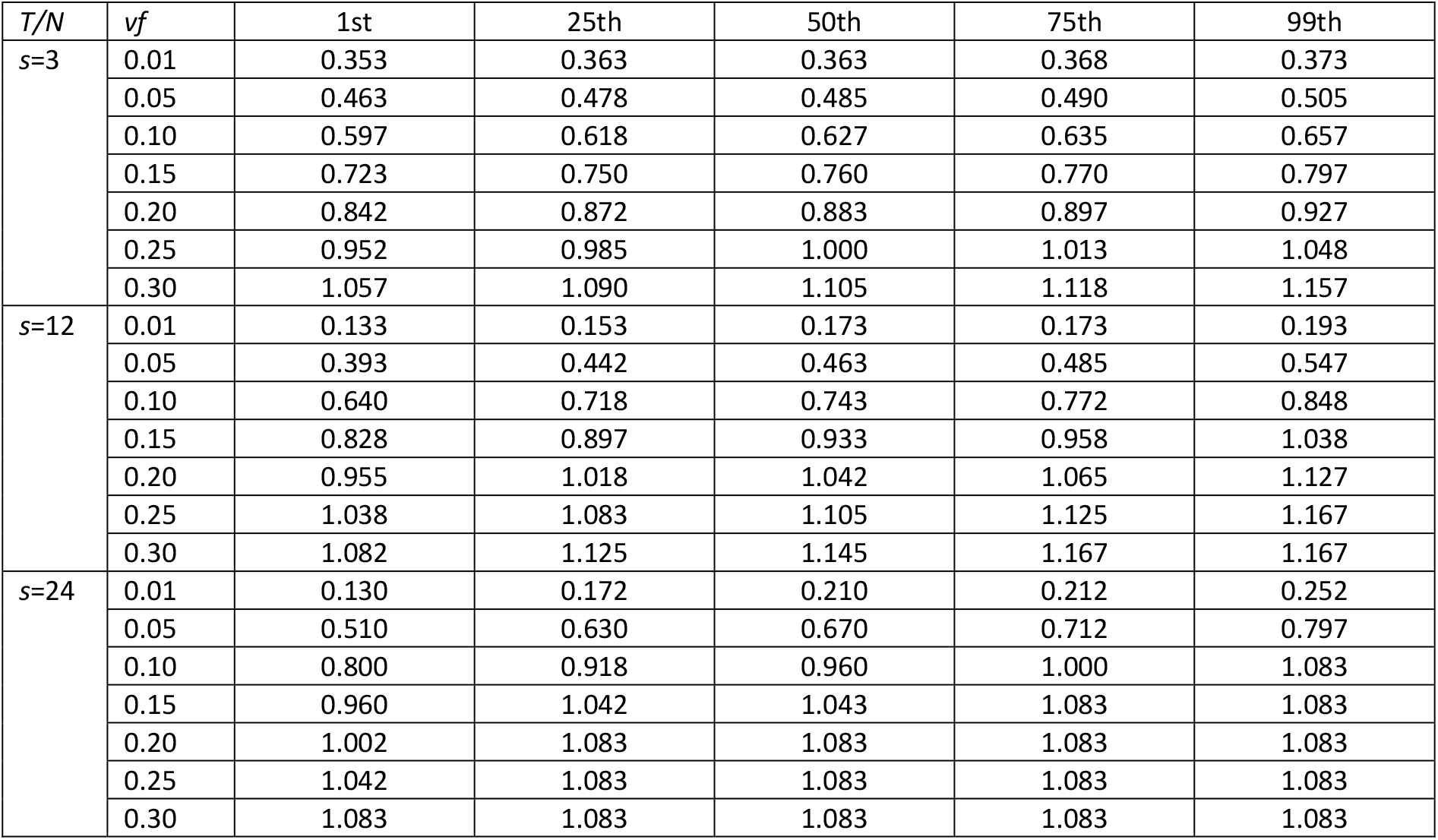
Random sequential pooling. Values of the 1st, the 25th, the 50th (median), the 75th, and the 99th percentiles of *T/N* obtained in the set of 5,000 simulations, for three values of the pool size (*s*=3, 12, 24) and some values of the virus frequency (*vf*=0.01, 0.10, 0.15, 0.20, 0.25, 0.30).

**Figure 4.**
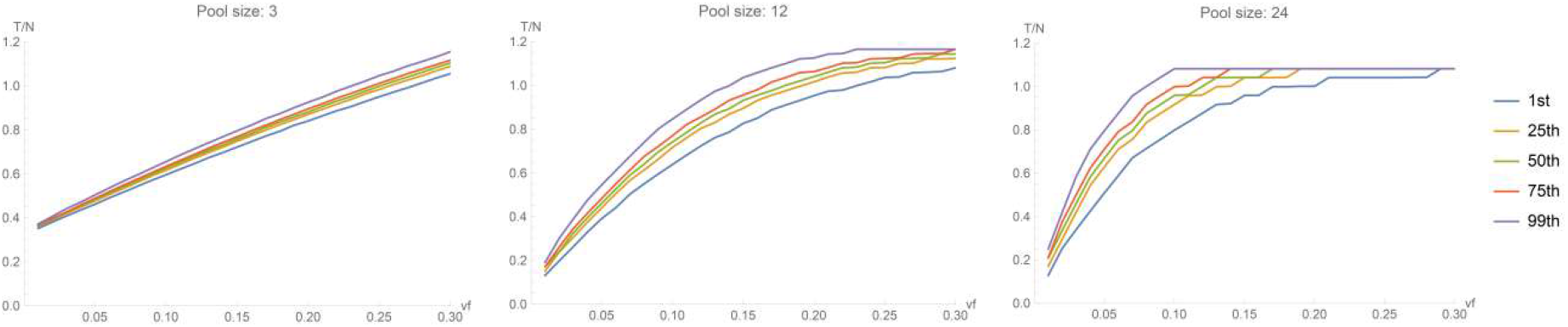
Random sequential pooling. The curves plotted represent the 1st, the 25th, the 50th (median), the 75th, and the 99th percentiles of *T/N* obtained in the set of 5,000 simulations, for three values of the pool size (*s*=3,12,24).

As mentioned in the Introduction, simple pooling has been recently proposed in Hogan et al [7]. We notice that their study does not provide efficiency results apart from their specific application, where pools of size 9 and 10 have been used and a very small *vf* has been reported (their value is even smaller than the smallest virus frequency assessed in our simulations). Figure 5 shows a comparison of a simple one-step pooling strategy with our two-step sequential procedure for different *vf* and *s* values. In this picture, the 25th, the 50th (median), and the 75th percentiles of *T/N* are shown. For very small pools (*s*=5), they are almost equivalent. But, as soon as *s* is slightly increased to sensible values (ranging from 8 to 20), the sequential two-step pooling shows a better performance up to *vf*=0.15. For bigger pools (*s*=24, 30), we observe the same result up to *vf* around 0.10. For higher frequency virus, both pooling strategies are counter-productive, as highlighted before for sequential pooling.

**Figure 5.**
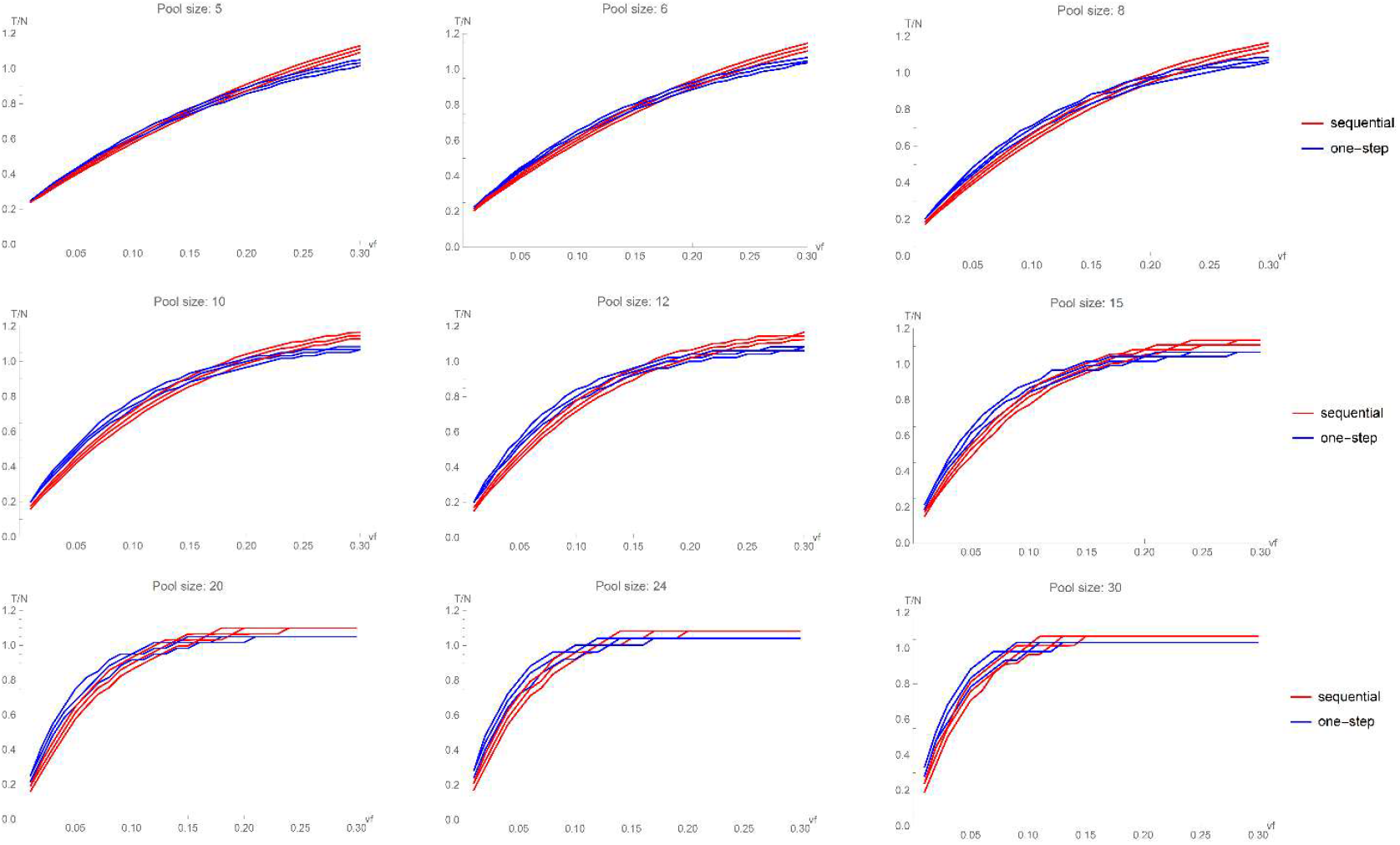
Random sequential pooling vs one-step pooling. The curves plotted represent the 25th, the 50th (median), and the 75th percentiles of *T/N* obtained in the set of 5,000 simulations, for 9 values of the pool size (*s*=5, 6, 8, 10, 12, 15, 20, 24, 30).

As explained, all the previous results have been obtained assuming a completely random assignment of subjects to the pools. Often, however, clinic and epidemiologic data about the subjects are available. If we could use these data to concentrate a portion of the positive subjects in the same horizontal pools, we would increase efficiency due to a higher number of negative pools at step B. In order to assess the savings of such an “informed pooling creation”, we extended our simulations to different settings. Imagine that, prior to the test we detect a certain number of subjects, say *x*, that we expect to be positive (according to epidemiological criteria). We create thus *x/s* horizontal pools, each of size s, with those subjects. The remaining *(N-x)* subjects are assigned to the remaining *(N-x)/s* horizontal pools. Should epidemiological criteria be perfect, all the *x* subjects turn out to be true positive and thus the first *x/s* pools are positive. At the same time, all the *(N-x)* subjects without prior indication of an infection, with perfect epidemiological criteria, would be true negative and thus their *(N-x)/s* horizontal pools would give a negative result. Of course, such an assumption is unrealistic and we expect that some of the *x* subjects suspected to be positive are true negative and also some of the *(N-x)* subjects suspected to be negative are true positive. Let us denote by α the fraction of the *vf·N* true positive subjects in the population that are correctly assigned to the initial pools. The remaining (1-α) fraction is undetected and it is wrongly assigned to the second part of the pools. Criteria with perfect performance in prior detection of positive subjects would result in α=1. In addition, let us denote by β the fraction of the (1-*vf)·N* true negative subjects in the population that are correctly assigned to the final pools. The remaining (1-β) fraction is wrongly assigned to the first part of the pools. Criteria with perfect performance in prior detection of negative subjects would result in β=1. For the same settings analysed in the random creation of the pools (*N*=600, *vf* from 0.01 to 0.30, and *s* from 2 to 300), we explored the performance of the sequential procedure for different values of α and β. In particular, we allowed α and β to vary in the set {0.5, 0.6, 0.7, 0.8}. When both α and β are equal to 0.5, criteria are essentially unreliable and our situation is equivalent to the random assignment setting above discussed.

Figure 6 shows the results of the simulations obtained for three values of the pool size (*s*=3,12,24) for different combinations of α and β (the plots for the remaining *s* values are shown in the Supplementary Material section, Figures S.2, S.3). Since there are many situations, to improve clarity, we plotted only the 50th percentile (the values for all the 5 percentiles are displayed in Tables 2a, 2b, 2c). Our aim is to compare the results of the number of tests required when swabs are randomly assigned to the pools with the number of tests required for different α and β values. As above mentioned, we started with α and β equal to 0.5, because this is substantially equivalent to uninformative prior criteria. As α and/or β increase, we observe that the number of required tests decreases, and this decrease is larger when the virus frequency is larger. When *vf* is below 5%, random pooling and informed pooling are almost equivalent. With a low *vf*, sequential random pooling was however already very performant, substantially decreasing the number of test with respect to separate individual tests. For larger *vf*, the curves corresponding to random assignment and informed pooling separate more and more. This entails that reliable informed pooling increases the performance of the pooling exactly when the situation is less favourable. For example, with a pool size equal to 12, with a random assignment, the median of *T/N* is equal to 1 when *vf* ≈ 0.18 (making random pooling application questionable). Conversely, if informed pooling is performed with α=β=0.8, at the same *vf*, the median of *T/N* is equal approximately to 0.73. With α=β=0.8, pooling is still efficient (*T/N<1)* even if the virus frequency approaches 30%. In summary, reliable informed pooling makes the performance path much more linear than we observed for random pooling, even if we use larger pools. That is, larger pools, besides providing substantial savings for low *vf*, ensure efficiency even for larger *vf* if epidemiological criteria provide reliable information.

**Table 2a.**
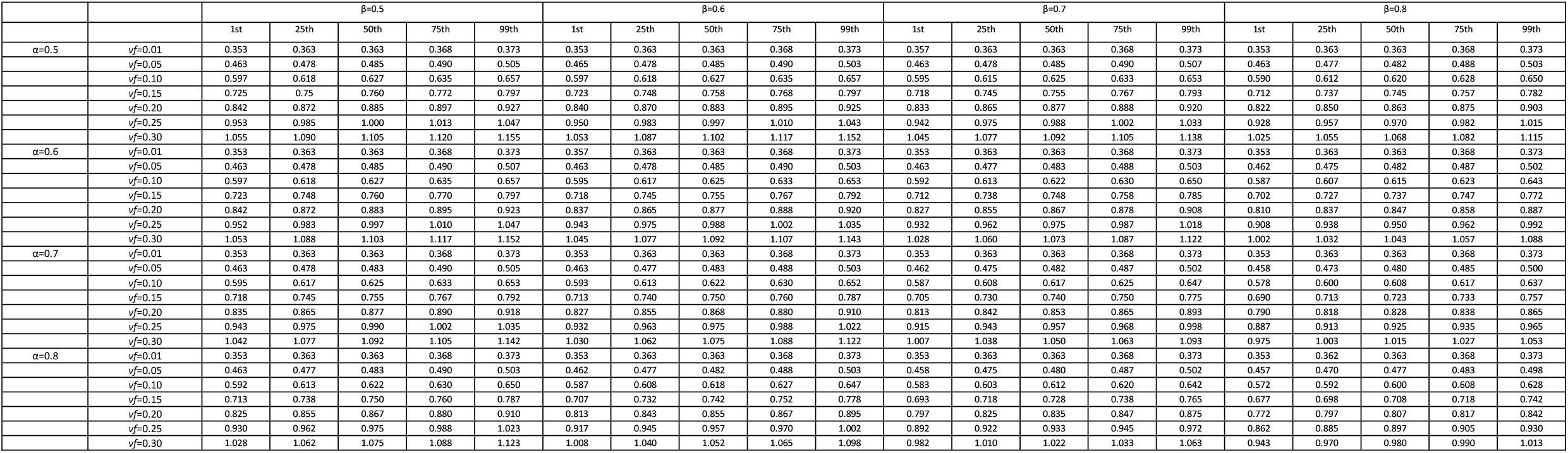
Informed sequential pooling, pool size *s*=3. Values of the 1st, the 25th, the 50th (median), the 75th, and the 99th percentiles of *T/N* obtained in the set of 5,000 simulations, for all combinations of α and β in {0.5, 0.6, 0.7, 0.8}, for some values of the virus frequency (*vf*=0.01, 0.10, 0.15, 0.20, 0.25, 0.30).

**Table 2b.**
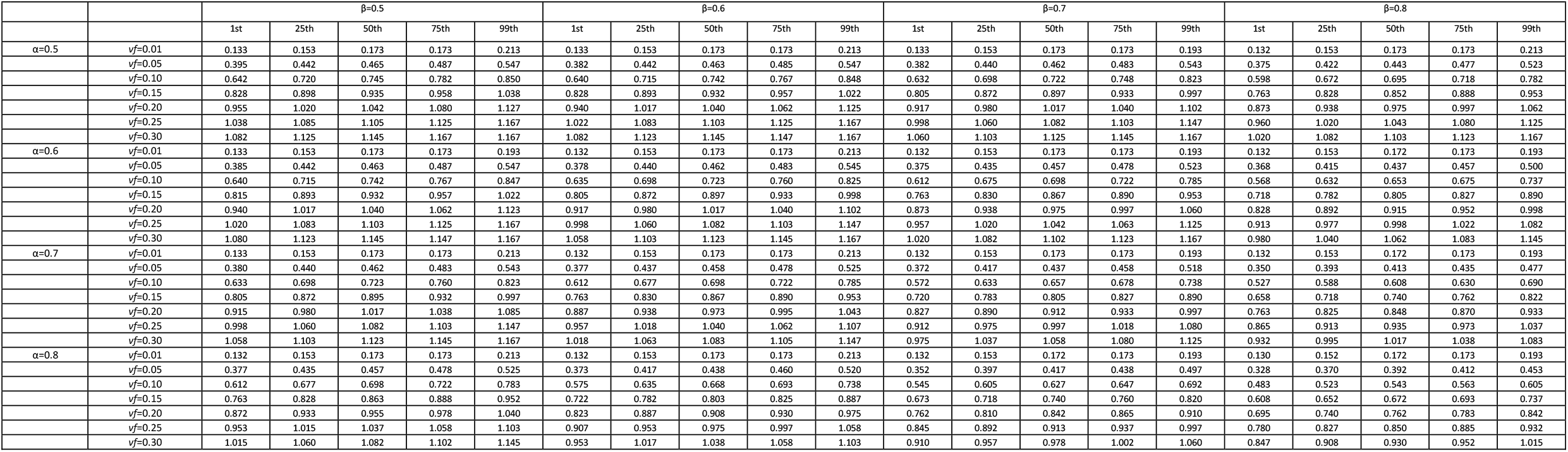
Informed sequential pooling, pool size *s*=12. Values of the 1st, the 25th, the 50th (median), the 75th, and the 99th percentiles of *T/N* obtained in the set of 5,000 simulations, for all combinations of α and β in {0.5, 0.6, 0.7, 0.8}, for some values of the virus frequency (*vf*=0.01, 0.10, 0.15, 0.20, 0.25, 0.30).

**Table 2c.**
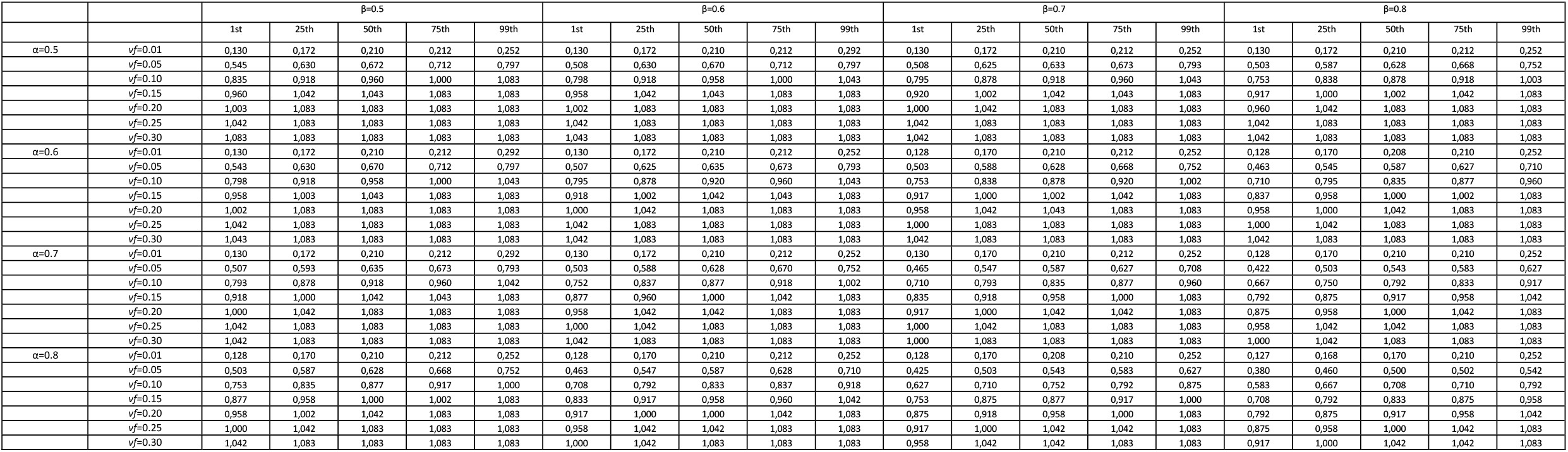
Informed sequential pooling, pool size *s*=24. Values of the 1st, the 25th, the 50th (median), the 75th, and the 99th percentiles of *T/N* obtained in the set of 5,000 simulations, for all combinations of α and β in {0.5, 0.6, 0.7, 0.8}, for some values of the virus frequency (*vf*=0.01, 0.10, 0.15, 0.20, 0.25, 0.30).

**Figure 6.**
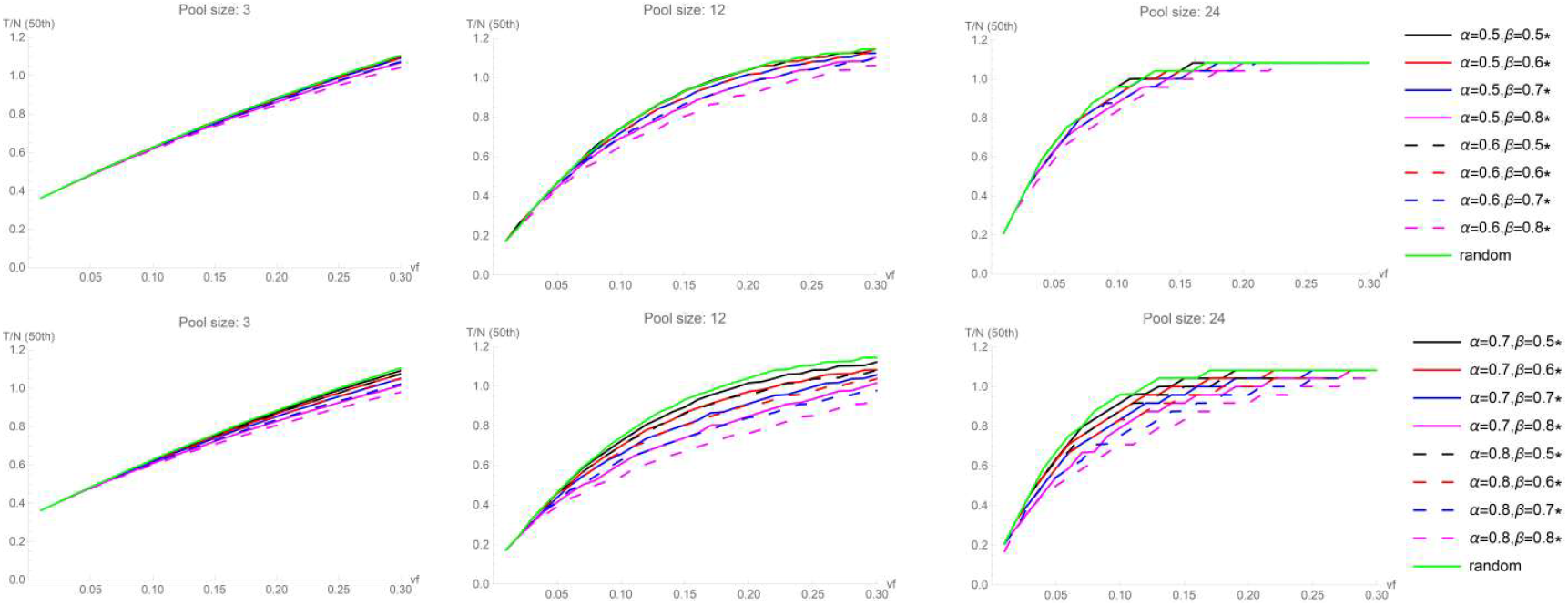
Informed sequential pooling. The curves plotted represent the 50th percentile (median) of *T/N* obtained in the set of 5,000 simulations, for three values of the pool size (*s*=3,12,24). The upper plots were obtained with α=0.5 and α=0.6, combined with β=0.5, 0.6, 0.7, 0.8. The lower plots were obtained with α=0.7 and α=0.8, combined with β=0.5, 0.6, 0.7, 0.8.

## Discussion

Every successful strategy in controlling SARS-CoV2 infection depends on the viral timely diagnosis. Hence, there is an urgent need for a systematic population screening at a scale mass. Currently, around the world, there is a plethora of different scenarios, which mainly depends on the spread of infection. Even in the same country, we can find very different situations: for example, it is likely that the asymptomatic population has a lower viral frequency than the symptomatic one or than who belongs from a category at risk, such as the health personnel. Moreover, in this variegated context, there are completely different economic situations, and the pooling strategy can become truly attractive for countries with fewer resources. The study published on JAMA [7] is certainly an excellent starting point to evaluate an alternative approach to individual analysis of swab sample for the RT-PRC based diagnosis of SARS-CoV2, but some appropriate considerations are needed. Firstly, it must be highlighted that in the JAMA study, 292 pools of 9 or 10 samples were created and two positive cases in a collection of 2888 samples were founded. The one-step pooling method gave excellent results because the frequency of the virus in the analyzed samples was extremely low (0.07%). Secondly, if it was possible to roughly estimate the frequency of the virus in the collection as lower than 5%, our data suggest to increase the pool size. Using a pool size of 24, for example, the screening of the 2888 samples would needed about 120 tests instead of 292.

Our most straightforward result is that the sequential pooling approach is more efficient than one-step pooling method. In addition, the informed version of the sequential pooling can further improve its performance, in particular for larger size pools and moderate to large virus frequency. Table 3 broadly describes practical suggestions to decide the pool size, *s*, according to rough assumptions about the virus frequency, both for random and informed sequential pooling. Larger pools ensure very large reduction in the number of tests when *vf* is small. Smaller pools may be a conservative approach when dealing with cohorts with heavier exposure. Finally, indications are also given to avoid the use of pooling when virus frequency is higher and random pooling would result in a waste of resources, since too many pools are expected to give a positive result.

**Table 3.** Summary of practical indications to pooling creation.

| Random sequential pooling | Informed sequential pooling ($\alpha, \beta \geq 0.7$) |
| --- | --- |
| If we can guess a $vf$ below 10%, pools with size from 10 to 15 can provide relevant savings in the number of tests. With $vf$ below 5%, even stronger savings can be obtained with pool size increased to 20 or 25. | If we can guess a $vf$ below 10%, very large pools, with size from 20 to 25 could substantially reduce the number of tests. Pools with size equal to 30 or 40 are a good strategy with $vf$ below 5%. |
| For situations when $vf$ may reach 10%-20% of the cohort, we can still have a moderate reduction in the number of tests, with pools of size between 5 and 8. | For situations when $vf$ may reach 10%-20% of the cohort, we can still have a relevant reduction in the number of tests, with pools of size between 12 and 20. |
| For situations when there is the risk of a $vf$ value above 20% of the cohort, pooling strategies should be avoided. | For situations when there is the risk of a $vf$ value above 20% of the cohort, a moderate reduction can be attained with pools of size about 12, to be further reduced to 8, if the $vf$ may exceed 25%. |

## Data Availability

All data are fully available without restriction.All relevant data are within the manuscript and its Supporting Information files. The corresponding author is

## Supplementary Material

In this Section, we list some Figures pertaining to the results of the simulations. In the main paper, only a selection of the plots could be included.

**Figure S.1.**
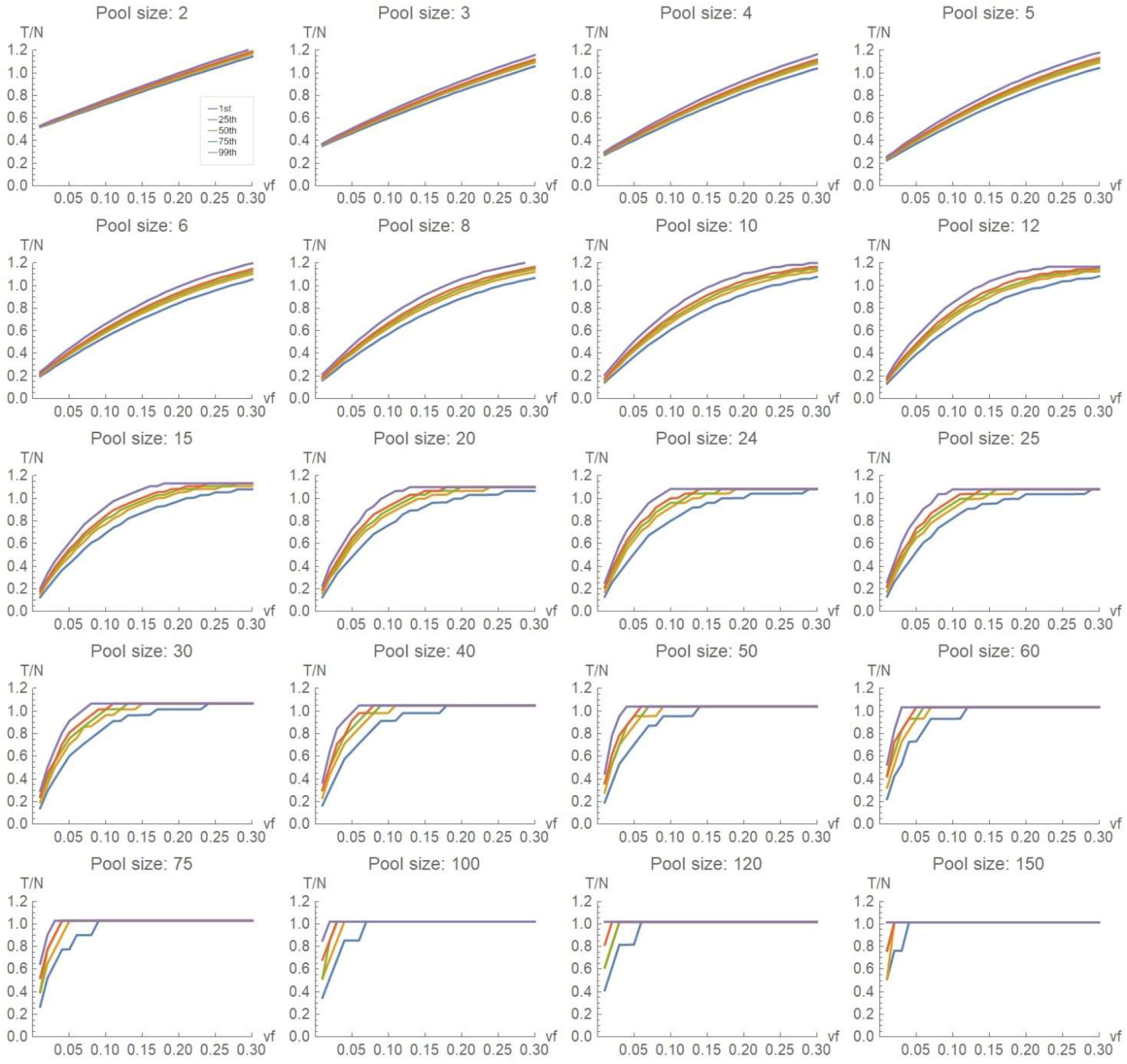
Random sequential pooling. The curves plotted represent the 1st, the 25th, the 50th (median), the 75th, and the 99th percentiles of *T/N* obtained in the set of 5,000 simulations, for a three values of the pool size *s* from 2 to 150.

**Figure S2.**
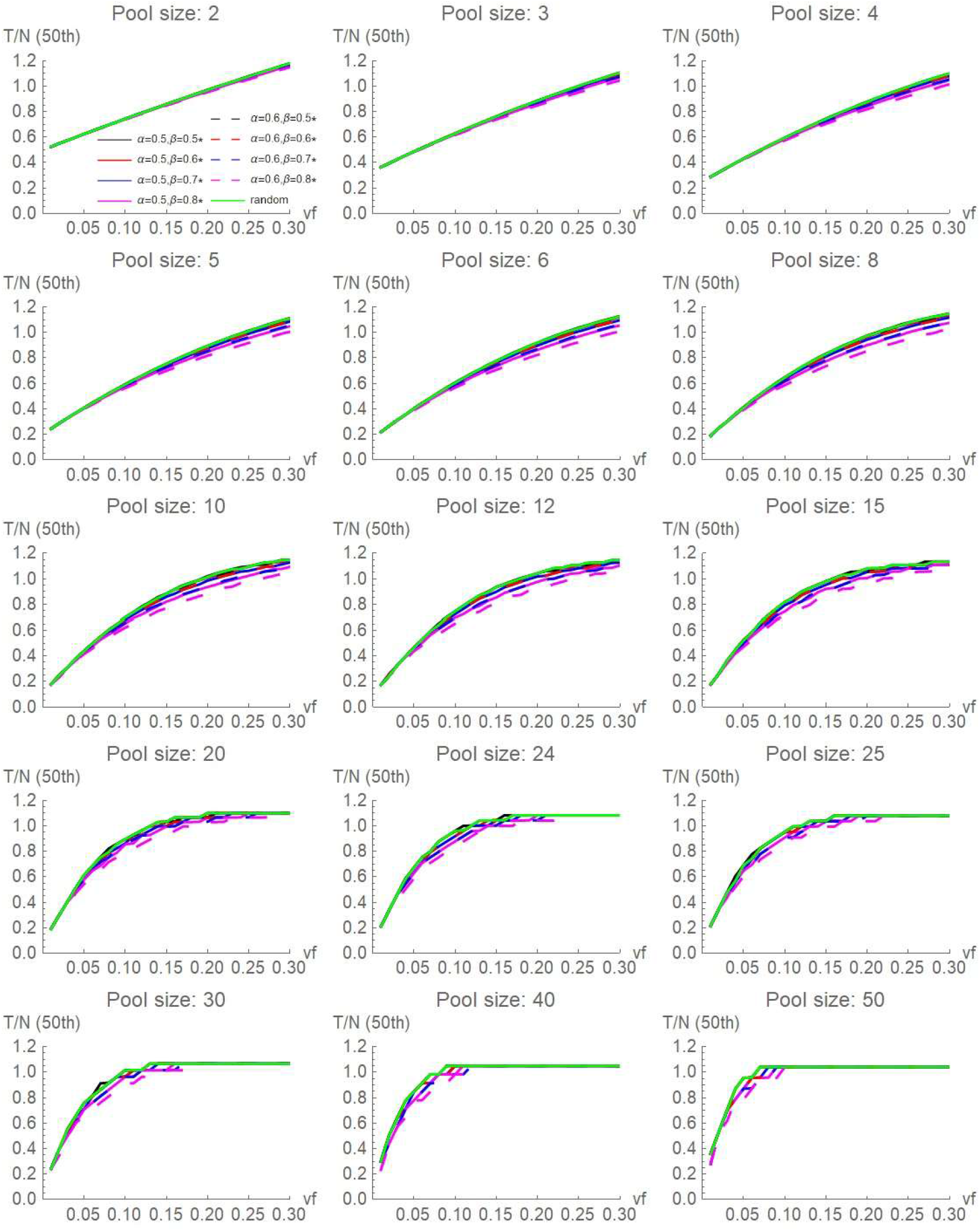
Informed sequential pooling. The curves plotted represent the 50th percentile (median) of *T/N* obtained in the set of 5,000 simulations, for values of the pool size *s* ranging from 2 to 50. The plots were obtained with α=0.5 and α=0.6, combined with β=0.5, 0.6, 0.7, 0.8.

**Figure S3.**
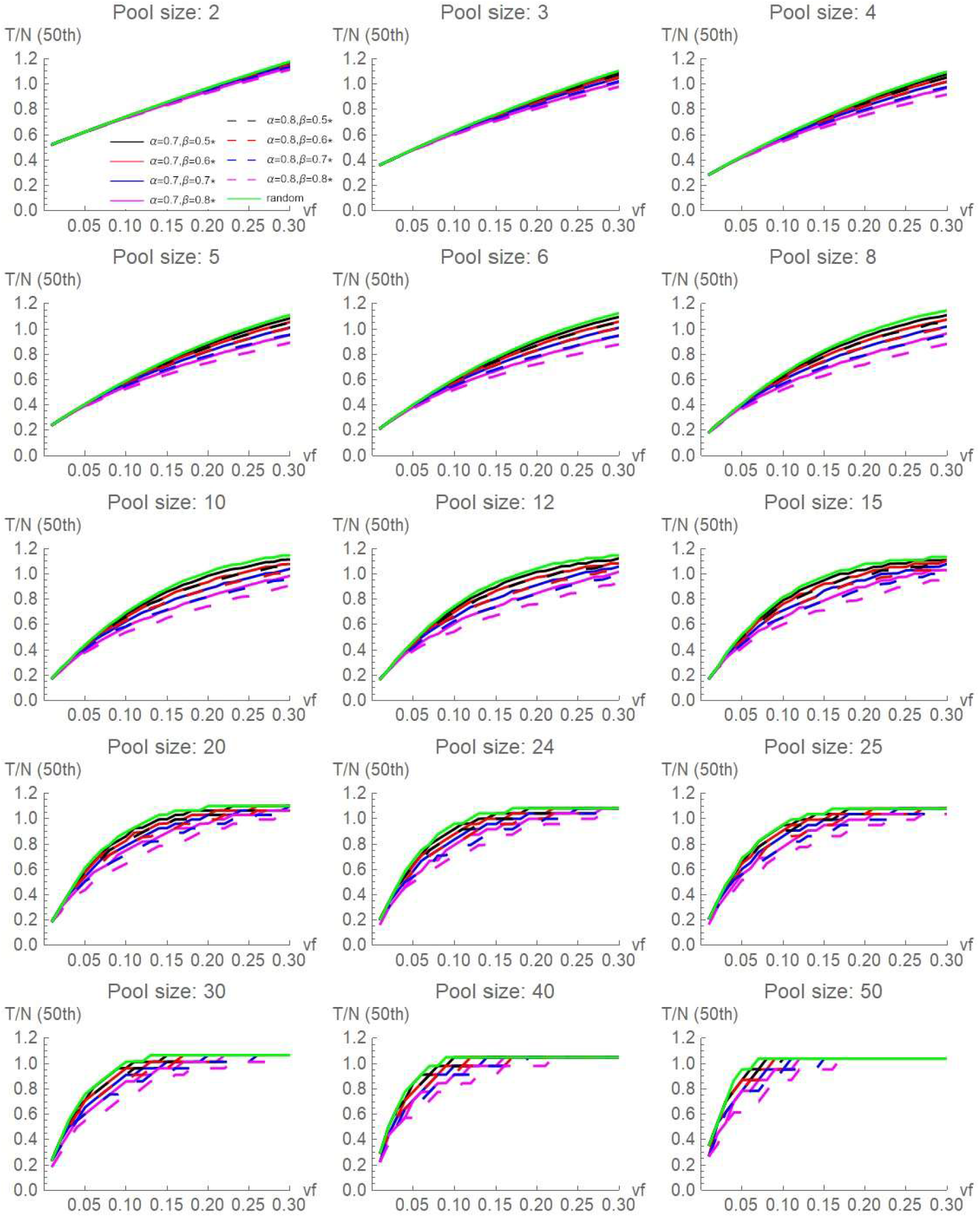
Informed sequential pooling. The curves plotted represent the 50th percentile (median) of *T/N* obtained in the set of 5,000 simulations, for values of the pool size *s* ranging from 2 to 50. The plots were obtained with α=0.7 and α=0.8, combined with β=0.5, 0.6, 0.7, 0.8.

## Footnotes

1 Results were obtained with Wolfram Mathematica 12.1.

2 The list of possible sizes *s* to split *N*=600 subjects is equal to {2,3,4,5,6,8,10,12,15,20,24,25,30,40,50,60,75,100,120, 150,200,300}.

## Notes

### Competing Interest Statement

The authors have declared no competing interest.

### Funding Statement

author

